# Nonlinear Markov Chain Modelling of the Novel Coronavirus (Covid-19) Pandemic

**DOI:** 10.1101/2020.04.21.20073668

**Authors:** Muammer Catak, Necati Duran

## Abstract

Almost all countries around the world are struggling against the novel coronavirus (Covid-19) pandemic. In this paper, a nonlinear Markov chains model is proposed in order to analyse and to understand the behaviour of the Covid-19 pandemic. The data from China was used to build up the presented model. Thereafter, the nonlinear Markov chain model is employed to estimate the daily new Covid-19 cases in some countries including Italy, Spain, France, UK, the USA, Germany, Turkey, and Kuwait. In addition, the correlation between the daily new Covid-19 cases and the daily number of deaths is examined.

## 1 Introduction

The novel coronavirus (Covid-19) pandemic has been spread all over the world [12]. According to Coronavirus disease 2019 (COVID-19) Situation Report-82 published by WHO, the total number of infected people approached to 2 million. However, in the same report WHO states that the true-number of the infected people might be higher than the number of the confirmed cases. Many researched have been published in order to analyse and to understand the dynamics of the Covid-19 pandemic [9, 13, 5, 14, 7, 10, 6]. Guiliani et al. [8] proposed a multivariate time-series linear model in order to understand and estimate the spatio-temporal diffusion of the Covid-19 pandemic in Northern Italy. It is stated that the proposed model is capable of to predict with an error of 3% compared to the late available data in Italy. Kucharski et al. [11] developed a mathematical model to understand the early dynamics of the transmission of the Covid-19 pandemic. They divided the entire population into four subsets as; susceptible, exposed, infectious, and removed.

In this study a mathematical model based on nonlinear Markov chains is proposed in order to estimate of the daily new Covid-19 cases. The paper s organized as follows: In Section 2, materials and methods employed in the proposed model is presented. Results obtained from the model are depicted in Section 3, and a brief discussion is given. Finally the paper is concluded in Section 4.

## 2 Materials and Method

Let *S*_*t*_ is a random process having countable number of states. If the process is in *i*^*th*^ state at time *t*, then it will be in *j*^*th*^ state at time *t* + 1 with the propability of *p*_*ij*_ defined as [4];

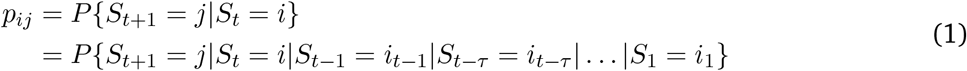

The state probability distribution vector *S*(*t*), the transition time step *τ*, and the transition matrix *P* are the main parameters will be used in a Markov chain model. In this study, the states are defined as *i*) 1 := Infected, *ii*) 2 := Recovered, *iii*) 3 := Dead. Thereafter, the nonlinear Markov cahin model can be expressed as [3];

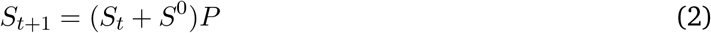

where *S*_*t*+1_ is the next state probability distribution, *S*_*t*_ is the current state probability distribution, *S*^0^ is the new infected distribution, and *P* = *P* (*t, S*_*t*_) is the Markovian transition matrix. The transition probabilities of the proposed nonlinear Markov chains model between the states are illustrated in Figure 1. Since the state-3 is an absorbing state, *p*_33_ is assigned as 1. The remaining entries of the transition matrix *P*, i.e *p*_*ij*_, have been calculated based on the confirmed daily number of Covid-19 cases and the daily test numbers of the corresponding countries.

The related website of John Hopkins University [2] has been used in order to get data in accordance with [1]. The number of the daily new cases, the number of Covid-19 test done, the number of the recovered cases, and the number of daily deaths have been used as the proposed model parameters affecting on the transition probabilities between the predefined states. The data obtained from China is employed to developed the model. The daily new Covid-19 cases and the corresponding estimation of the proposed nonlinear Markov chain model is illustrated in Figure 2. In China, the confirmed new daily Covid-19 cases on 11 February 2020, which is the 21^*st*^ day after the first case was reported on 22^*nd*^ January 2020, is assumed as a deviation from the model due to the test policy.

**Figure 1:**
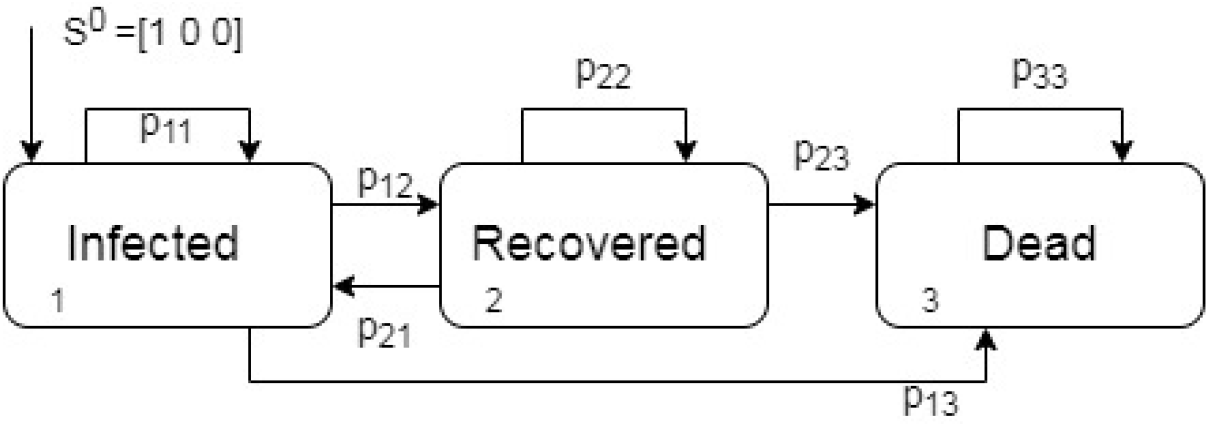
Illustration of the transition probabilities of the proposed model

**Figure 2:**
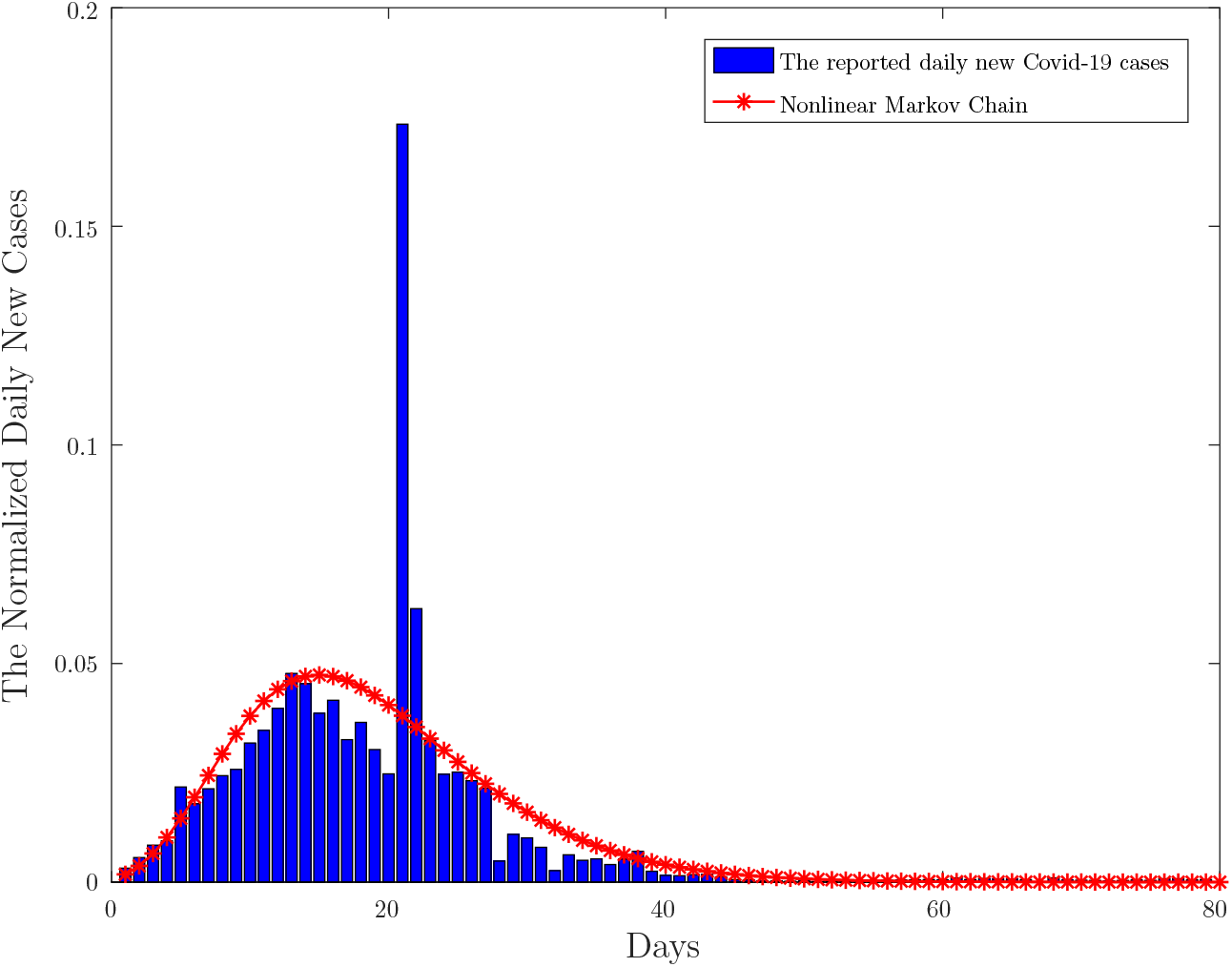
The daily reported new Covid-19 cases and the prediction of the proposed nonlnear Markov chain model

## 3 Results and Discussion

In this section, the proposed nonlinear Markov chain model is employed in order to estimate the daily new Covid-19 cases in Italy, Spain, France, UK, Germany, USA, Turkey, and Kuwait. Please note that, it is assumed that the Covid-19 pandemic is over if the number of the daily new Covid-19 cases are less than 50 considering 3 consecutive days apart from Kuwait where the threshold was taken as 10 instead of 50 considering the population of State of Kuwait.

The daily new Covid-19 cases of the aforementioned countries are shown in Figure 3 and Figure 4. The first Covid-19 case was reported in Italy on 22 February 2020. It seems the Covid-19 pandemic has reached its peak after almost 40 days from the first case was announced, then it started to decrease the number of the Covid-19 cases daily bases. The proposed model estimates that the Covid-19 pandemic will be over about the first week of May 2020 results in more than 200 thousands of the infected person.

**Figure 3:**
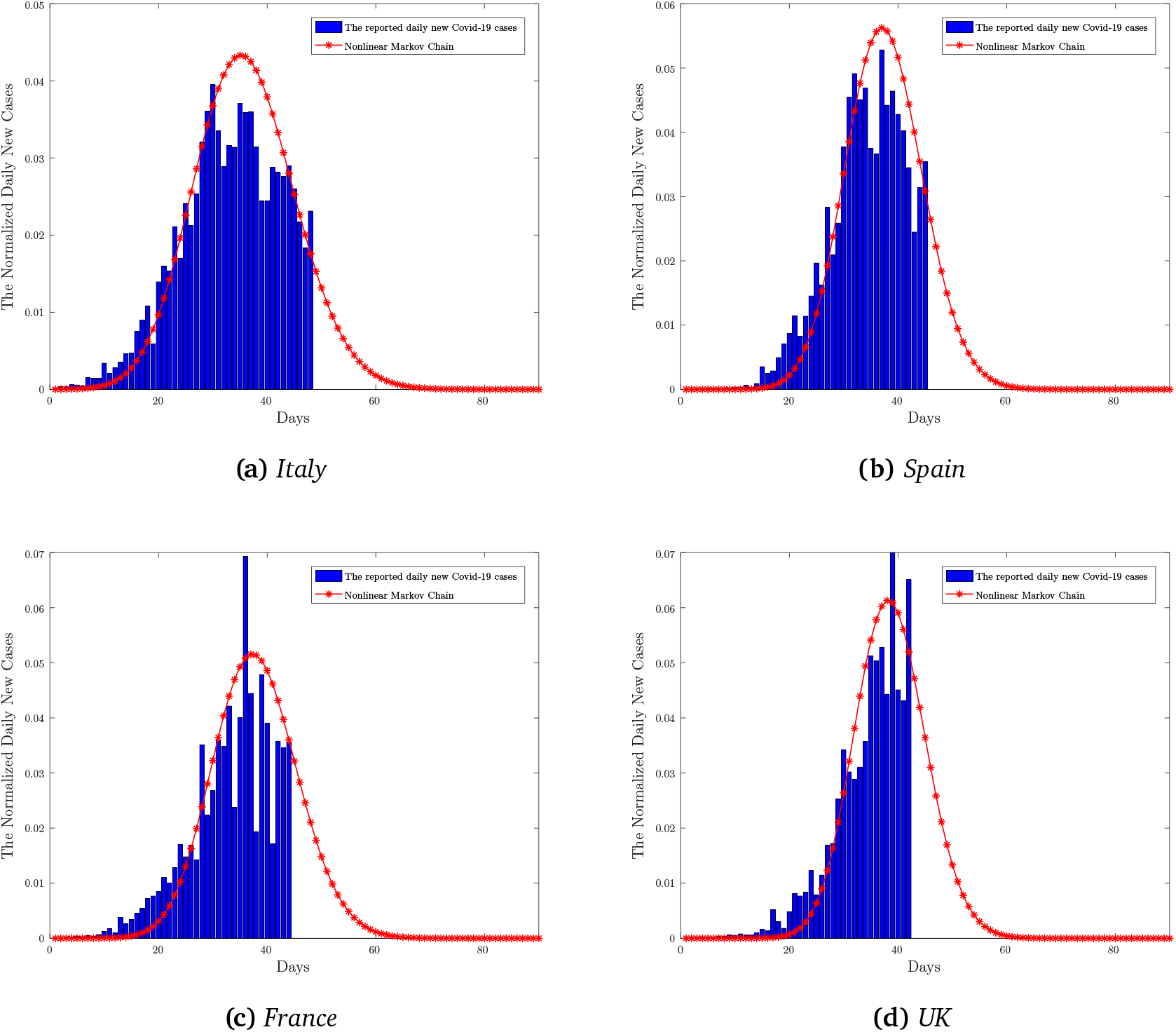
The daily number of new Covid-19 cases

**Figure 4:**
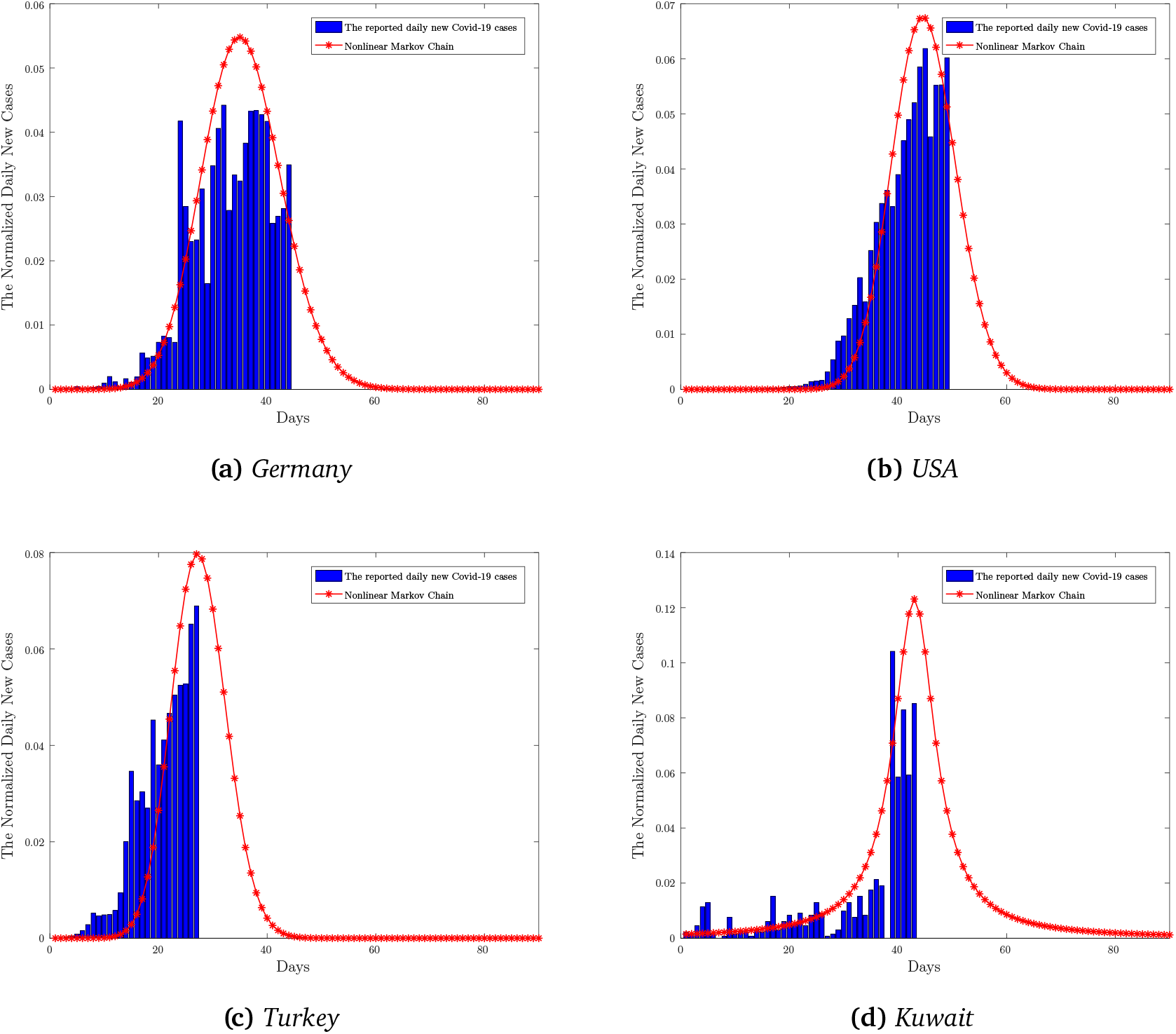
The daily number of new Covid-19 cases

A detailed statistical information about the related countries are given in Table 1. In general, the first cases of the Covid-19 were reported about end February, 2020. Thereafter, the Covid-19 pandemic reached its peak values in the first half of April, 2020. Finally, the model estimates that the Covid-19 pandemic will be over before the first week of May, 2020.

**Table 1:** Estimations of the proposed Nonlinear Markov chains model for some countries

| Country | First Case Reported | Peak Date | Estimation of the End Date | Estimation of the Total number of Infected |
| --- | --- | --- | --- | --- |
| Italy | 22 February | 1 April | 4 May | 206.000 |
| Spain | 25 February | 5 April | 2 May | 228.000 |
| France | 26 February | 6 April | 5 May | 202.000 |
| UK | 28 February | 8 April | 3 May | 169.000 |
| Germany | 26 February | 4 April | 28 April | 174.000 |
| USA | 21 February | 4 April | 8 May | 975.000 |
| Turkey | 11 March | 12 April | 7 May | 138.000 |
| Kuwait | 24 February | 8 April | 10 May | 2.650 |

Moreover, the correlation between the daily new Covid-19 cases and the daily number of deaths are analysed considering the corresponding countries. The number of *lag* – *k* where the cross correlation of the functions reaches its maximum are given in Table 2.

**Table 2:** lag – k parameters of the countries

|  | China | Italy | Spain | France | UK | Germany | USA | Turkey |
| --- | --- | --- | --- | --- | --- | --- | --- | --- |
| $lag - k$ | 6 | 1 | 1 | 3 | 1 | 5 | 1 | 1 |

The cross correlation between two sequences can be explained as;

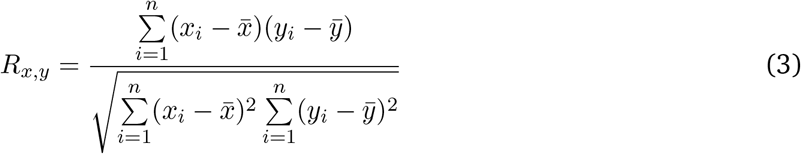

where 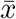 and 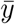 are the expected values of the sequences of *x* and *y*, respectively.

The value of *lag* – *k* can be considered as the time-shift between the daily number of the Covid-19 cases and the daily number of deaths. Therefore, the increases in *lag* – *k* parameter shows that the corresponding country has detected the infected people earlier. Moreover, it might be a positive sign of the health system in such a country. There are only 2 reported dead cases in Kuwait up to now, hence State of Kuwait is not listed in Table 2.

## 4 Conclusion

In this paper, an nonlinear Markov chain model is proposed in order to analyse and to understand the novel coronavirus (Covid-19) pandemic. The data obtained from China was used to build up the presented model. Then, the proposed nonlinear Markov chain model is employed to predict the daily new Covid-19 cases among some countries. According to the results, the proposed model can efficiently capture the general behaviour the Covid-19 pandemic dispersion. Please note that, all the reported data including the daily number of new Covid-19 cases, the daily number of deaths, the number of the daily Covid-19 test are all assumed as correct. The model might need to be refined when a more reliable and detailed data is available.

## Data Availability

All data is available on the related websites given in the manuscript.

https://www.worldometers.info/coronavirus/

https://coronavirus.jhu.edu/map.html

## Notes

### Competing Interest Statement

The authors have declared no competing interest.

### Funding Statement

No funding

